# Immune-sensitization to *Mycobacterium tuberculosis* Among Young Children With and Without Tuberculosis

**DOI:** 10.1101/2025.01.16.25320625

**Authors:** Jesús Gutierrez, LaShaunda L. Malone, Mitchka Mohammadi, John Mukisa, Michael Atuhairwe, Simon Peter G. Mwesigwa, Salome Athieno, Sharon Buwule, Faith Ameda, Sophie Kiyingi, Ezekiel Mupere, Catherine M. Stein, Christina L. Lancioni

**Author notes:** Contributed equally as co-senior authors (Catherine M. Stein, Christina L. Lancioni). **Address correspondence to: Christina Lancioni**, MD. Department of Pediatrics, Oregon Health and Science University, Portland, OR. [ ].

## Abstract

**Background:** Identification of young children with *Mycobacterium tuberculosis* (*Mtb*)-infection is critical to curb Tuberculosis (TB)-related pediatric morbidity and mortality. The optimal test to identify young children with evidence of *Mtb*-infection remains controversial.

**Methods:** Using a TB household contact (HHC) study design among 130 Ugandan children less than 5 years with established *Mtb*-exposure, we compared the usefulness of the tuberculin skin test (TST) and QuantiFERON Gold Plus (QFT-Plus) to identify children with evidence for *Mtb*-sensitization. We conducted univariate analysis to compare findings between children with and without TB disease, and performed a logistic regression model to estimate the odds of TB. We performed a sensitivity analysis by stratifying results by age (< 2 years vs. 2-5 years). Finally, we compared results of the QFT-Plus TB tube 1 and TB tube 2 to establish concordance.

**Results:** A 5 mm TST threshold identified the most children with evidence of *Mtb*-sensitization; this result was most pronounced in children with TB. Moreover, the odds of TB were 2 times higher [aOR: 2.09 (CI: 1.02 – 4.37)] among children with a positive TST. The QFT-Plus’ TB tube 1 and TB tube 2 results were highly correlated.

**Conclusions:** TST identified more TB-exposed young children with evidence of *Mtb-*immune-sensitization, when compared to QFT-Plus. These findings are highly relevant for children who are TB HHCs in endemic settings, and most at risk for TB following an exposure. We recommend that TST testing continue to be performed to assess for *Mtb*-sensitization in young children.

## BACKGROUND

Over 1 million children develop Tuberculosis (TB) annually, with close to 250,000 pediatric deaths attributed to TB every year (1). Due to difficulties in diagnosing TB in children and limitations in public health resources in many TB-endemic settings, most children who die from TB never receive treatment (2–4). Identification of children at risk of TB due to recent *Mycobacterium tuberculosis* (*Mtb*)-infection, and improvements in diagnostic approaches for pediatric TB, are global public health priorities (1, 4).

Progression to TB disease following *Mtb*-infection can be prevented with TB preventive therapy (TPT) (5, 6). Therefore, prompt identification of young, *Mtb*-infected children, remains the most effective tool to curb TB-related pediatric morbidity and mortality. Currently, there is no reference standard test for asymptomatic *Mtb*-infection. Rather, evidence of immunologic memory to *Mtb* (referred to as *Mtb*-sensitization) is considered a surrogate for *Mtb*-infection and to identify individuals who would most benefit from TPT. Similarly, evidence of *Mtb*-sensitization is often utilized to support a clinical diagnosis of TB in young children undergoing evaluation for TB disease, as microbiologic confirmation of TB remains extremely challenging in this age group (2). Both the tuberculin skin test (TST) and interferon-gamma release assays (IGRAs) can be used to identify individuals with evidence of *Mtb*-sensitization (2, 7–10). Both tests serve as indicators of immunologic-memory to mycobacterial antigens, and neither assay can reliably distinguish between individuals with asymptomatic or subclinical infection and those with active TB disease. A primary limitation of TST is the potential for nonspecific responses following immunization with bacilli Calmette-Guérin (BCG) vaccine and/or exposure to environmental mycobacteria (11–13). Thus, thresholds to determine a positive TST are controversial and dependent on factors such as age, HIV-status, nutritional status, country of origin, and TB-exposure history (9).

IGRAs, such as the QuantiFERON Gold Plus (QFT-Plus), do not cross-react with antigens present in BCG vaccine or environmental mycobacteria and are more specific than TST for *Mtb*-sensitization (7, 8). Unlike the original QuantiFERON Gold test, the QFT-Plus includes a second antigen tube that captures CD8+ T-cell responses intended to increase the assay’s sensitivity (14), although recent studies have not supported this conclusion (14, 15). The threshold to define a positive IGRA does not consider age, immunocompromising conditions, or TB exposure. For young children, particularly those under 2 years who are most at risk for rapid progression to TB following an exposure, the optimal test to identify those with *Mtb*-sensitization remains controversial (9, 14, 16), and discordance between TST and IGRA results have been widely reported (17). Identification of young children with evidence of *Mtb*-sensitization is critical to identify asymptomatic children who would most benefit from TPT (18), and in clinical decision making to diagnose TB in unwell children with known TB exposure (19). Here, we hypothesized that in a population of young children with extensive exposure to TB in the home, TST and IGRA testing would perform similiarly to identify individuals with evidence for Mtb-sensitization. Using a pediatric TB household contact study design among Ugandan children less than 5 years with and without TB, we compared the usefulness of the TST and QFT-Plus to identify children with evidence of *Mtb*-sensitization.

## METHODS

### Setting

This study was conducted in Kampala, Uganda, a highly TB endemic setting (20).

### Population

The study population included 130 children ages < 5 years who are household contacts (HHCs) of an adult or adolescent with confirmed pulmonary TB disease (index case). A HHC is defined as a child living in the same home of the index case for ≥ 7 consecutive days in the past three months. All index cases had symptomatic pulmonary TB disease, confirmed by positive sputum AFB culture (detected on either liquid or solid media) or *MTB*/RIF GeneXpert® Ultra assay. The parent(s) or guardian(s) of HHCs < 5 years were contacted to undergo informed consent within 28 days of the index case’s TB diagnosis. HHCs were excluded if they had a prior history of being treated for TB, were currently receiving treatment for TB, or were expected to be unavailable for the 12-month follow-up period. The study protocol was approved by the Makerere University School of Biomedical Sciences Research Ethics Committee, the Uganda National Council on Science and Technology, and the institutional review board at University Hospitals Cleveland Medical Center. Written informed consent was obtained from the parent or guardian of each HHC.

### Design

This was a cross-sectional examination of data obtained during evaluation and follow-up of pediatric HHC enrolled in a 12-month longitudinal study designed to understand the immunobiolgy of *Mtb*-infection and TB disease in young children. At study entry, each child underwent HIV testing, QFT-Plus testing, and TST placement (interpreted within 2-3 days); maternal HIV status during pregnancy and/or breastfeeding was established. Based on HIV results, each child was characterized as HIV-unexposed/uninfected (HUU), HIV-exposed/uninfected (HEU), or a child living-with-HIV (CLWH). QFT-Plus was performed in a CAP-certified laboratory and results interpreted according to the manufacturer’s recommendations. The TST was performed by the Mantoux method (5 tuberculin units per 0.1 ml of purified protein derivative, Tubersol; Connaught Laboratories Limited, Willowdale, Ontario Canada). We considered TST positivity using two different cutoffs. First, following WHO guidelines (21), 10 mm of induration indicated a positive TST, except for CLWH or severe wasting (defined by length/height-for-weight Z-score ≤ -3) when a 5 mm cutoff was used. We derived anthropometric z-scores from WHO child growth standards (22). Second, we applied a cutoff of 5 mm or greater of induration for all participants, regardless of HIV or nutritional status, per Center for Disease Control (CDC) guidelines for pediatric TB HHC (9). Each HHC underwent review of medical history and TB symptoms, complete physical examination (including assessment for disseminated and/or extrapulmonary TB), nutritional assessment via anthropometry (height/length, weight, BMI, and upper arm circumference), 2-view chest radiographs (CXR; interpreted by radiologist; Supplemental Figure 1), and collection of two induced sputum samples for AFB smear, culture, and Xpert Ultra testing, as well as an individual risk assessment. This risk assessment was administered to compute an epidemiologic risk score (ERS) that quantified degree of *Mtb* exposure. This score ranges from 0 to 10 with a higher score conferring an increased risk. This standard questionnaire included information on risk factors such as degree of contact with the index case, relationship to the index case, index symptoms, and index case clinical characteristics (23). To estimate the duration of household TB exposure among enrolled children, the number of days of TB-related symptoms among each index case was also collected (Supplemental Table 1).

Based on the results of their baseline evaluation, each HHC was assigned to one of two initial cohorts by a study physician using standardized criteria (16): HHCs without signs or symptoms of TB disease, and a negative diagnostic evaluation for TB disease, regardless of TST and QFT-Plus results, were considered an asymptomatic exposure cohort. HHCs in the asymptomatic exposure cohort were offered TPT using 6 months of INH treatment, per WHO recommendations for all pediatric TB HHC < 5 yo (24). HHCs with at least 1 sign or symptom of TB disease and/or CXR findings consistent with pulmonary TB, and/or a positive microbiologic evaluation for TB disease, were considered to have TB disease. Children with TB diseases received 6 months of TB treatment according to Ugandan national guidelines. Following completion of the 12-month study, children received a final cohort assignment of asymptomtic *Mtb-*exposure (PedsAS) or TB disease (PedTB) using a standardized consensus review process and review of all clinical and microbiologic data. Children with a positive induced sputum assay for *Mtb* (by either AFB cultures or Xpert Ultra) were considered to have confirmed TB disease, whereas children meeting clinical criteria for TB with negative sputum studies were considered to have unconfirmed TB (19) (Supp Table 2).

### Statistical Analyses

Analysis included the first 130 pediatric HHC to complete the 12-month study; *a priori* power calculations were not performed. We conducted univariate analysis to compare final TB classification cohorts (PedAS and PedTB) using demographic information, ERS, HIV classification, BCG status, quantitative and categorical QFT-Plus results, quantitative and categorical (using both 5 mm and 10 mm cut-offs) TST results, and anthropometric z-scores. Comparisons were performed using Fisher’s exact test and chi-square test for categorical variables, and student’s t-test and Mann-Whitney *U* test for continuous variables. Significance was assessed using a 0.05 alpha cut-off. We used the results of this univariate analysis to create a logistic regression model to estimate the odds of being diagnosed with TB. Second, we assessed the concordance between TST and QFT-Plus categorical results among all study participants using a McNemar’s test and the Kappa statistic and then performed a sensitivity analysis by stratifying the results by age (< 2 years vs. 2-5 years) and final cohort classification (PedAS vs. PedTB). Finally, we compared the results of TB tube 1 (TB.1) and TB tube 2 (TB.2) of the QFT-Plus test using Spearman’s correlation to evaluate the concordance of these results and to assess if the use of TB tube 2 added value in identification of *Mtb* immune-sensitization. All analyses were performed using R software (25). Data collection and management for this paper was performed using OpenClinica open-source software (version 3.16. Copyright OpenClinica LLC and collaborators, Waltham, MA, USA, www.OpenClinica.com).

## RESULTS

### Demographic and clinical characteristics, diagnostic results, and TB classification among study participants

Children were heavily exposed to TB in their homes, with a mean ERS of 7.0 among the entire population (Table 1). The presence of prolonged TB symptoms (cough, fever, sputum production, night sweat, hemoptysis, and weight loss) was common among index cases, with a median duration of 60 days for cough and sputum production (Supplemental Table 1). Among all children, 31.5% were QFT-Plus positive, 39.2% were TST positive using a 10 mm threshold, and 49.2% were TST positive using a 5 mm threshold (Table 1). Among 130 HHCs, we classified 75 as PedAS and 55 as PedTB. The majority of 55 HHCs classified as PedTB (90.9%) had findings consistent with TB on CXR (Table 2). Notably, all 5 children with normal CXR findings in the PedsTB cohort had microbiologically confirmed TB. One-third of children with a final classification of PedTB presented with symptoms consistent with TB (Table 2), and five had confirmed TB (Table 2). During their baseline evaluation, several children who did not received a final classification of PedTB were noted to have abnormal CXR and/or symptoms consistent with TB such as cough and fever. Among these 13 children, 9 were treated with an empiric course of antibiotics at their initial study visit with resolution of symptoms and 4 children were provided symptomtic care (eg., anti-pyretics, anti-histamines, steroids, zinc) for suspected viral and/or asthma related illnesses with improvment reported during their follow-up.

**Table 1:** Demographic characteristics, QFT-Plus, and TST results of study participants with and without TB.

|  | Full Cohort | PedAS | PedTB | p-value (test) |
| --- | --- | --- | --- | --- |
| **n** | 130 | 75 | 55 |  |
| **Age (months)** | 27.5 [±16.2] | 25.9 [±16.2] | 29.6 [±16.2] | 0.20 (t-test) |
| **Sex = male** | 73 (56.2) | 42 (56.0) | 31 (56.4) | 1.00 ($\chi^2$) |
| **Epidemiologic Risk Score (range: 0-10)** | 7 [6.0-8.0] | 7 [6.0-9.0] | 7 [6.0-8.0] | 0.12 (MWU) |
| **HIV classification** |  |  |  | 0.86 (Fisher's) |
| HIV unexposed uninfected (HUU) | 87 (66.9) | 48 (64.0) | 39 (70.9) |  |
| HIV exposed uninfected (HEU) | 19 (14.6) | 12 (16.0) | 7 (12.7) |  |
| Child living with HIV | 4 (3.1) | 2 (2.7) | 2 (3.7) |  |
| HIV exposure unknown | 19 (14.6) | 12 (16.0) | 7 (12.7) |  |
| HIV result pending<sup>a</sup> | 1 (0.8) | 1 (1.3) | 0 (0) |  |
| **IGRA (QFT-Plus) = positive<sup>b</sup>** | 41 (31.5) | 21 (28.0) | 20 (36.4) | 0.34 ($\chi^2$) |
| **Mean quantitative IGRA result<sup>c</sup>** | 0.01 [0-2.47] | 0.02 [0-1.77] | 0.03 [0-4.96] | 0.75 (MWU) |
| **Mean quantitative IGRA result<sup>c</sup> in children with positive IGRA (n=41)** | 6.59 [2.72-9.22] | 5.34 [1.98-9.51] | 6.89 [4.63-7.97] | 0.82 (MWU) |
| **TST = positive (10 mm)** | 51 (39.2) | 25 (33.3) | 26 (47.3) | 0.15 ($\chi^2$) |
| **TST = positive (5 mm)** | 64 (49.2) | 31 (39.7) | 33 (61.1) | **0.02\*** ($\chi^2$) |
| **Quantitative TST result** | 3.4 [0-14.9] | 0 [0-13.5] | 8.6 [0-16.3] | **0.02\*** (MWU) |
| **BCG scar present** | 112 (86.2) | 64 (85.3) | 48 (87.3) | 0.80 (Fisher's) |

Counts (percentages), means [± standard deviation], or median [quartiles]
$\chi^2$ : Chi-squared test
MWU: Mann–Whitney *U* test
QFT-Plus: QuantiFERON-TB Gold Plus
□ Child will undergo testing after breastfeeding is complete
<sup>b</sup>Does not include 5 participants with indeterminate results
<sup>c</sup>Calculated as (QFT-Plus tube 1 + QFT-Plus tube 2) / 2
**\* Statistically significant at $p < 0.05$**

**Table 2:** Summary of diagnostic evaluation for TB among study participants.

|  | **PedAS** | **PedTB** |
| --- | --- | --- |
| **n** | **75** | **55** |
| **Chest X-ray (CXR)** |  |  |
| Normal | 70 (93.3%) | 5 (9.1%) |
| Moderately advanced disease<sup>a</sup> | 3 (4.0 %) | 50 (90.9%) |
| Unknown | 2 (2.7%) | 0 (0%) |
| **TB symptoms** |  |  |
| No symptoms | 62 (82.7%) | 36 (65.5%) |
| At least one symptom | 13 (17.3%) | 19 (34.5%) |
| **Extra-pulmonary TB symptoms** |  |  |
| No symptoms | 75 (100%) | 55 (100%) |
| At least one symptom | 0 (0%) | 0 (0%) |
| **Xpert Ultra result** |  |  |
| Negative | 75 (100%) | 51 (92.7%) |
| Positive | 0 (0%) | 3 (5.5%) |
| Unknown | 0 (0%) | 1 (1.8%) |
| **Smear Microscopy result** |  |  |
| Negative | 74 (98.7%) | 53 (96.4%) |
| Positive | 0 (0%) | 2 (3.6%) |
| Unknown | 1 (1.3%) | 0 (0%) |
| **Culture result** |  |  |
| Negative | 75 (100%) | 50 (90.9%) |
| Positive | 0 (0%) | 5 (9.1%) |

<sup>a</sup>Moderately advanced disease - disease may be present in one or both lungs; the total extent must not be more than the following:
- (a) scattered lesions of slight to moderate density may not involve more than total volume of one lung or the equivalent volume of both lungs. - (b) dense, confluent lesions may not involve more than 1/3 of the volume of one lung. - (c) the total diameter of cavity(ies) may not be greater than 4 cm.

When comparing the PedAS and PedTB cohorts, we found no statistically significant differences in age, sex, ERS, HIV exposure or infection, BCG status or anthropometric measurements (Table 1 and Supplemental Table 3). Five children (3.7%) had indeterminate QFT-Plus results and were excluded from subsequent analysis; indeterminate results were due to failure of mitogen to elicit a response (N=3) or a high background response (N=2). Using the recommended threshold for interpretation, QFT-Plus positivity rate was 31.5% across the entire cohort. There were no significant differences in categorical or quantitative QFT-Plus results between PedAS and PedTB cohorts, including when quantitative values were compared only among those children with a positive qualitative QFT-Plus result (Table 1). Notably, the PedTB cohort had a higher quantitative TST result (0 mm vs 8.6 mm, *p*=0.02) and a greater rate of TST positivity with the 5 mm cutoff, when compared to the PedAS cohort (61.1% vs. 39.7%, respectively, *p*=0.02). When using the 10 mm cutoff, the TST positivity rate was still greater in the PedTB cohort versus the PedAS cohort (47.3% vs. 33%, respectively, *p*=0.15), but this difference did not reach statistical significance (Table 1).

### Concordance between TST and QFT-Plus among study participants

Among all children, concordance between TST and QFT-Plus categorical results was higher with the 10 mm versus 5 mm threshold (Cohen’s κ: 0.74 vs. 0.59; Supplemental Tables 4 & 7)). Concordance between tests was higher among all children ages 2-5 years compared to all children younger than 2 years for both 10 mm (Cohen’s κ: 0.83 vs. 0.60, respectively) and 5 mm (Cohen’s κ: 0.72 vs 0.42, respectively) thresholds (Supplemental Tables 4 & 7). Among children classified as PedAS, concordance between TST and QFT-Plus categorical results was high among children ages 2-5 years using either a 10mm or 5 mm cutoff (Cohen’s κ: 0.88 and 0.83, respectively; Supplemental Tables 5 & 8). Among children classified as PedAS who were younger than 2 years, concordance between TST and QFT-Plus categorical results varied when using a 10mm vs 5 mm cutoff (Cohen’s κ: 0.72 and 0.55, respectively; Supplemental Tables 5 & 8). Among children classified as PedTB, concordance between TST and QFT-Plus categorical results varied in both age groups when using a 10mm vs 5 mm cutoff . Specificially, among children ages 2-5 years with TB, concordance between tests using a 10mm or 5 mm cutoff was Cohen’s κ= 0.75 versus and 0.56, respectively (Supplemtal Tables 6 & 9). Among children classified as PedTB who were younger than 2 years, concordance between tests was low when using a 10mm or 5 mm cutoff (Cohen’s κ= 0.39 versus Cohen’s κ= 0.20, respectively). (Supplemental Tables 6 & 9).

### Relationship between age and assessments for *Mtb*-sensitization among children with and without TB

Next, we explored the relationship between age and the positivity rate for the TST and QFT-Plus (Table 3). Among children ages 2-5 years, 50% of those classified as TB (PedTB) were QFT-Plus positive, and TST positivity was 56.3% versus 67.7% (10 mm versus 5 mm thresholds, respectively). Conversely, QFT-Plus and TST positivity rates were below 36% in those without TB (PedAS). Among children < 2 years, only 20% of the children diagnosed with TB demonstrated a positive QFT-Plus (Table 3). The TST positivity rate among these younger children with TB was 34.8% when using a 10 mm threshold and 52.2% when using a 5 mm threshold.

**Table 3.** Assessment of *Mtb* immune-sensitization among children with and without TB stratified by age.

|  |  | Under 2 years of age (n=57) |  |  | 2 to 5 years of age (n=73) |  |  |
| --- | --- | --- | --- | --- | --- | --- | --- |
|  |  | PedAS | PedTB |  | PedAS | PedTB |  |
| **IGRA<sup>a</sup>**<br>**(QFT-Plus)** | Positive | 8 (24.2%) | 4 (20%) | $p=0.98$ | 13 (35.6%) | 16 (50%) | $p=0.21$ |
|  | Negative | 25 (75.8%) | 16 (80%) |  | 27 (67.5%) | 16 (50%) |  |
| **TST (10 mm cutoff)** | Positive | 13 (38.2%) | 8 (34.8%) | $p=1.00$ | 12 (29.3%) | 18 (56.3%) | $p=0.04^*$ |
|  | Negative | 21 (61.8%) | 15 (65.2%) |  | 29 (70.7%) | 14 (43.8%) |  |
| **TST (5 mm cutoff)** | Positive | 16 (47.1%) | 12 (52.2%) | $p=0.91$ | 15 (35.7%) | 21 (67.7%) | $p=0.01^*$ |
|  | Negative | 18 (52.9%) | 11 (47.8%) |  | 27 (64.3%) | 10 (32.3%) |  |

QFT-Plus: QuantiFERON-TB Gold Plus
<sup>a</sup>Does not include 5 children with indeterminate QFT-Plus results
\*Statistically significant

### Role of TST in predicting TB

Next, we assessed the value of tests of *Mtb* immune-sensitization in predicting TB disease. Based on univariate analysis (Table 1), both the 5 mm TST categorical result and quantitative TST result could be included as predictors in our logistic regression model, with equivalent model fit statistics. Due to collinearity, we decided to include the categorical TST variable only while excluding the quantitative TST variable and the categorical QFT-Plus variable. Finally, we adjusted the model for age (months), sex, HIV, and BCG status (Supplemental Table 10).

As shown in Table 4, the odds of TB disease were approximately 2 times higher when the child had a positive TST using the 5 mm cutoff (*p* = 0.04). Using the same model, we re-evaluated the odds of TB disease when applying the 10 mm cutoff. Here, the odds of TB disease were not significantly different based on TST result.

**Table 4:**
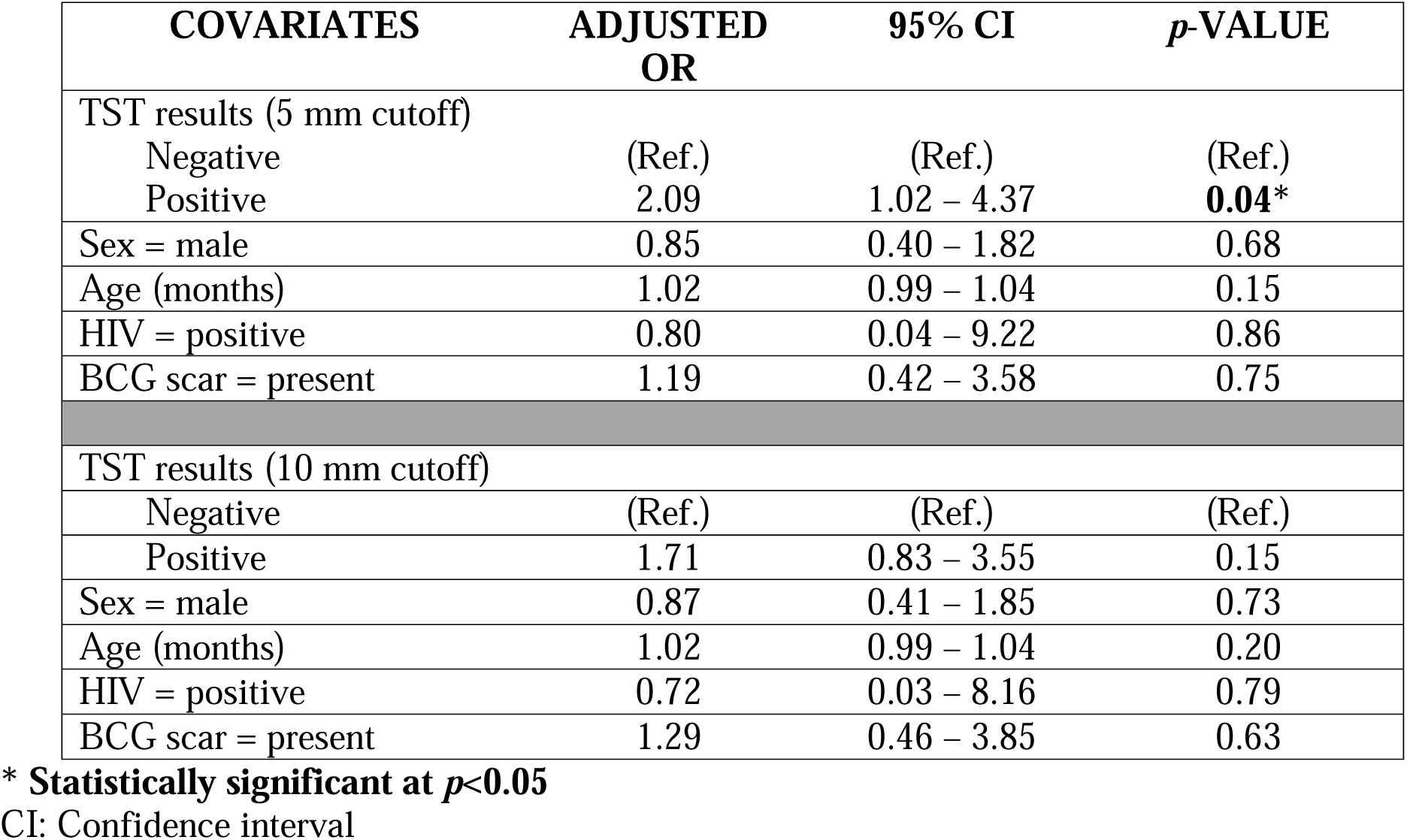
Logistic regression models demonstrating odds of TB disease using 5 mm and 10 mm TST thresholds.

### Contribution of QFT-Plus Tube 1 versus Tube 2 in identification of children with *Mtb*-immune sensitization

We examined the role of QFT-Plus Tube TB.1 and Tube TB.2 in identification of children with *Mtb* immune-sensitization. Among all participants, there was not a significant difference in median production of IFN-gamma between Tube TB.1 [median 0.01 IU/ml (IQR, 0-1.85)] and Tube TB.2 [median 0.02 IU/ml (IQR, 0-3.010-3.01]), as shown in Figure 1 and Supplemental Table 11. Furthermore, we demonstrated significant correlation between Tube TB.1 and Tube TB.2 (R=0.82; *p*<0.001). Only three children had discordant QFT-Plus Tube TB.1 and Tube TB.2 results, as circled in red in Figure 2 (Supplemental Table 12). Similarly, among the 41 participants with a positive QFT-Plus result, no significant differences in the median production of IFN-gamma between Tube TB.1 [median 6.12 IU/ml (IQR, 2.68-9.01)] and Tube TB.2 [median 6.14 IU/ml (IQR, 3.82-9.92)] were found. The correlation between Tube TB.1 and Tube TB.2 (R=0.86; *p*<0.001) also remained strong.

**Figure 1:**
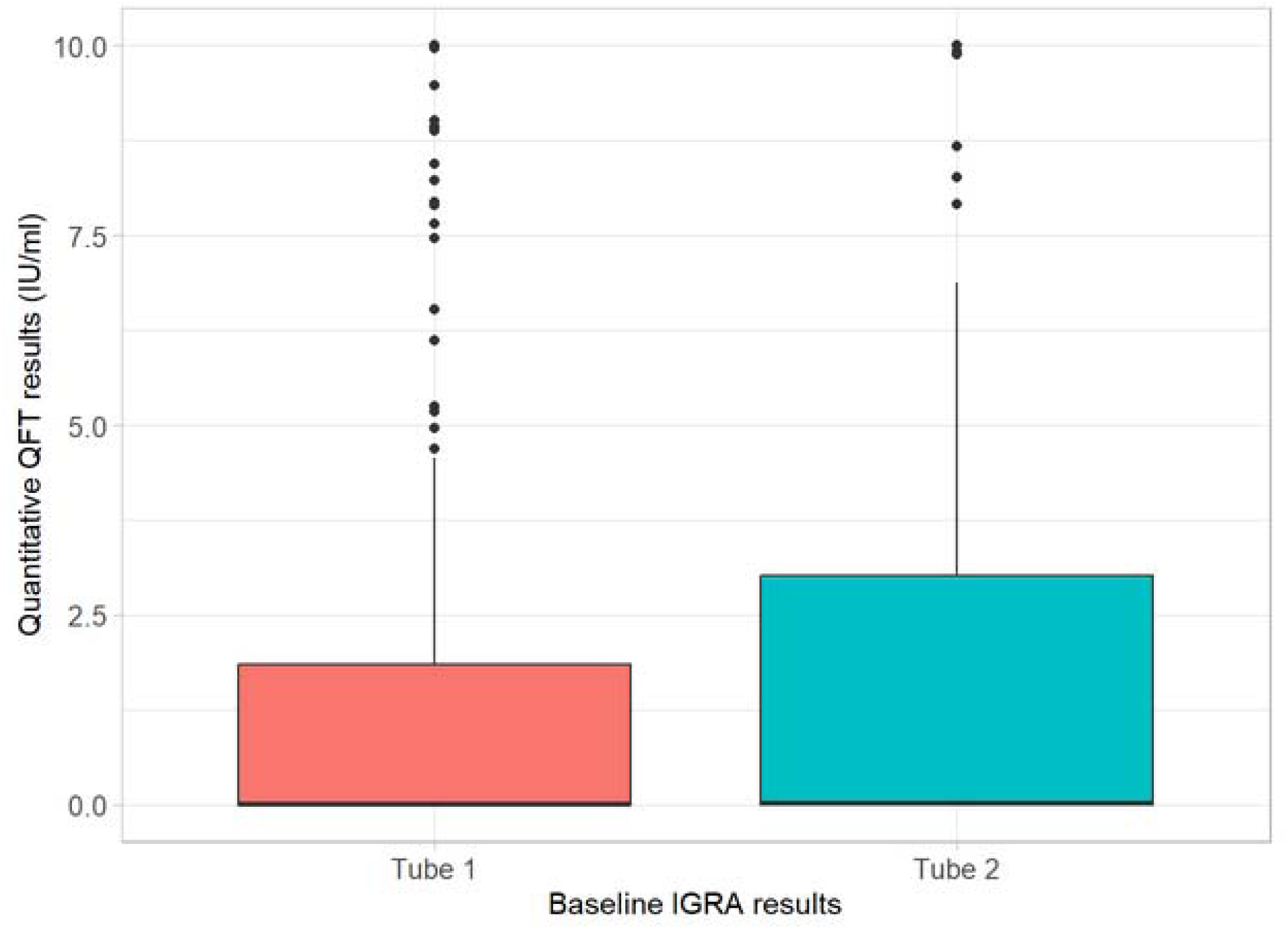
Comparison of Tube TB.1 and Tube TB.2 results of the QuantiFERON Gold TB-Plus assay. There were no differences when comparing the results of Tube TB.1 (median: 0.01 [IQR, 0-1.85]) and Tube TB.2 (median: 0.02 [IQR, 0-3.01], *p*-value=0.31. This comparison does not include 5 participants who had inderterminate results.

**Figure 2:**
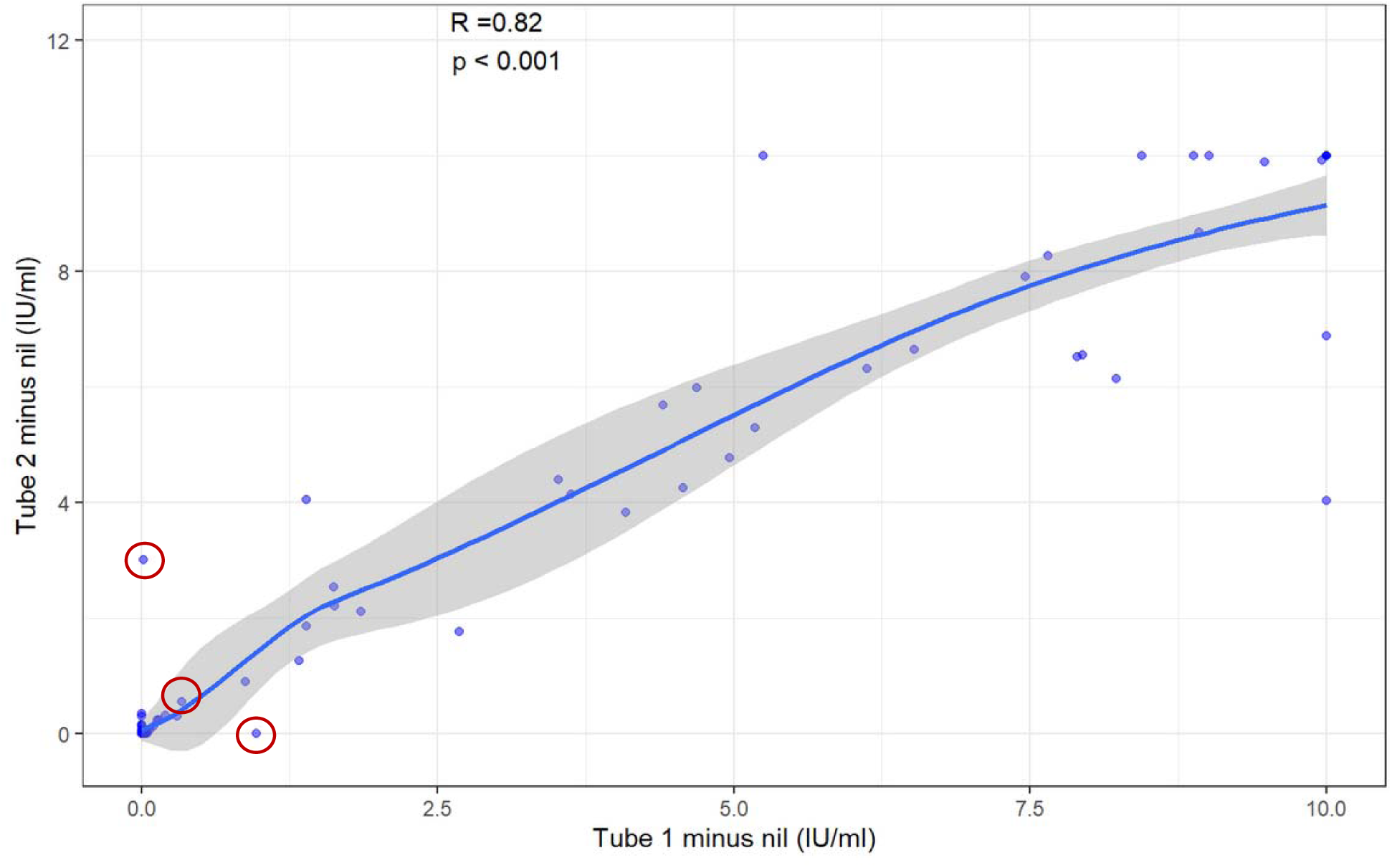
Correlation between quantitative QFT-Plus Tube TB.1 and Tube TB.2 results. Using the Spearman’s correlation coefficient (rho), we compared the results of TB.1 and TB.2. The results of TB.1 and TB.2 were strongly and positively correlated to each other based on a rho of 0.82 (*p*=<0.001). This analysis did not include five participants with indeterminate results.

## DISCUSSION

Identification of young children with evidence for *Mtb*-immune sensitization is critical to identify those who would most benefit from TPT, and to support a clinical diagnosis of TB among TB-exposed children with concerning signs and symptoms. Here, we employed a TB HHC study that included all children under 5 years living with an individual with microbiologically-confirmed pulmonary TB, to compare the capacity of TST and QFT-Plus testing to identify children with evidence of *Mtb*-sensitization. We found that despite the high ERS observed in this population of children, and prolonged cough and sputum production among index cases indicating extensive household TB exposure, TST and QFT-Plus testing demonstrated evidence of *Mtb*-sensitization in less than 50% of all exposed children. Overall, a 5 mm TST threshold identified the most children with evidence of *Mtb*-sensitization. Although neither the TST nor currently available IGRAs can reliably delineate between individuals with *Mtb* infection from those with TB disease, a positive result on either test is commonly used to support a clinical diagnosis of pediatric TB. Thus, it is notable that the enhanced performance of the TST was most pronounced in young children with TB.

Young children less than 5 years old are uniquely vulnerable to develop TB following recent *Mtb* infection, with progression to TB within 24 months of exposure in up to 20% of children (18, 26, 27). Young age is also associated with more severe clinical manifestations of TB and substantially higher mortality rates (27, 28). Therefore, the identification of young children at risk of TB disease due to recent *Mtb* infection, and improvement of diagnostic approaches for pediatric TB, remain global public health priorities (1, 4). Unfortunately, our findings suggest that neither the TST nor QFT-Plus are reliable indicators of *Mtb*-sensitization in a population of young Ugandan children with extensive TB exposure, particularly those under 2 years old. Our findings are consistent with previous studies where both tests had low sensitivity in young children with TB (29–33) or suspected disease (32–34). Moreover, our findings emphasize the limitations of immune-based diagnostics, such as IGRAs, in young children. Although introduction of a second tube containing *Mtb*-specific peptides (TB.2) in the QFT-Plus was anticipated to enhance detection of immune sensitization to *Mtb*, we found results to be highly correlated between TB.1 and TB.2, as reported in other studies in children (14, 15). We hypothesize that immune-sensitization to *Mtb*-infection in young children is not sufficiently identified using quantification of interferon-gamma production alone in response to *Mtb*-specific peptides. Several studies among asymptomatic Mtb-exposed children, and children being evaluated for TB disease, support that quantifying production of non-IFN-gamma, alternative cytokines, in response to Mtb-specific antigens is a promising approach to identifiy children with Mtb-sensitization and potentially delineate between infection and disease states (35–37).

IGRAs have become the preferred test for detection of *Mtb*-sensitization in well-resourced settings, particularly among individuals 5 years and older (9). In these populations, the enhanced specificity of IGRAs compared to TST can reduce unnecessary exposure to TPT among TST-positive BCG-vaccinated individuals (9, 38). In addition, several studies in young children suggest that BCG-vaccination is associated with a positive TST among individuals without risk factors for *Mtb*-exposure, and this association wanes with increasing age (39–42). One large study performed in South Africa demonstrated that IGRA results are more highly correlated with measures of TB exposure using the ERS as compared to TST positivity (43). Moreover, among South African children 2-5 years, the quantitative IFN-gamma response was strongly associated with TB disease status (44). In this current cohort of young Ugandan children, we did not observe either of these findings.

Concerns about false-positive TST results among BCG-vaccinated young children is a driving factor behind the WHO’s recommendation to apply a 10 mm threshold for positivity among children-without-HIV-infection and non-wasted children known to be exposed to TB (16). In the United States, however, where BCG-vaccination is not given, the CDC and American Academy of Pediatrics recommends a threshold of 5 mm for children with known TB exposure (9, 45). In TB endemic settings such as Uganda, WHO guidelines for TST interpretation are followed. However, our findings suggest that application of a 5 mm threshold for all TB HHCs under 5 years would substantially increase the identification of children with evidence for *Mtb*-sensitization, particularly those being evaluated for TB disease. Among asymptomatic, TB exposed young children, a TST threshold of 5 mm could be utilized to prioritize children for TPT when universal treatment is not feasible.

Our study has several limitations. Firstly, we did not have access to a sufficient number of BCG-unvaccinated, TB HHCs under 5 years, to assess associations between BCG-vaccination and TST results using 5 mm and 10 mm thresholds. We also did not have access to a similarly aged population of non-TB exposed Ugandan children to assess the specificity of TST versus QFT-Plus for *Mtb-*sensitization. As is expected among children treated for TB disease, the majority of children did not demonstrate microbiologically confirmed disease, despite testing two induced sputum samples for *Mtb* using AFB smear, AFB liquid and solid cultures, and Xpert Ultra (46–48). Sputum induction was selected as it has been shown to be safe, practical, and have similar yields for disease confirmation in young children as compared to first-morning gastric aspirates (49–51). We also note that TST results were available in real-time, whereas QFT-Plus results were delayed, and this could have biased an initial assignment of TST-positive children to the TB disease cohort. However, our final cohort assignments (completed following a 12-month period of observation) demonstrated that all children classified as having TB disease had either an abnormal CXR suggestive of pulmonary TB, microbiologic-confirmed disease, and/or signs and symptoms of TB disease; no child was classified as having TB disease solely upon the basis of their exposure history and a positive TST. Finally, TST and QFT-Plus testing was performed at study entry and not repeated, and we cannot rule-out that some children may have been in the window-period prior to development of an *Mtb*-specific adaptive immune responses. However, given that children were living in homes where index cases reported 1-2 months of TB symptoms, it is unlikely that repeat testing would have identified additional children with evidence of immune-sensitization.

## CONCLUSIONS

Overall, in a population of young Ugandan children with extensive household exposure to TB, the TST identified more children with evidence for *Mtb*-immune sensitization than QFT-Plus testing. The findings from this study are highly relevant for young children who are TB HHCs in an endemic setting with high rates of BCG-vaccination, limited resources to evaluate for *Mtb*-infection and TB disease, and poor access to TPT. As children under 5 years are extremely vulnerable to severe TB disease and remain the most challenging population to diagnose, we recommend that TST testing continue to be performed to assess for *Mtb*-sensitization in this age group, and that a 5 mm threshold be applied to those with known *Mtb*-exposure.

## Data Availability

All data produced in the present study are available upon reasonable request to the authors

## LIST OF ABBREVIATIONS

*Mtb*: *Mycobacterium tuberculosis*
TB: tuberculosis
HCC: household contact
TST: tuberculin skin test
IGRA: interferon-gamma release essay
QFT-Plus: QuantiFERON Gold Plus
WHO: World Health Organiztion
BCG: bacilli Calmete-Guérin
HUU: HIV-unexposed/uninfected
HEU: HIV-exposed/uninfected
CLWH: a child living-with-HIV
PedTB: diagnosed with TB
PedAS: not diagnosed with TB
ERS: Epidemiologic risk score
FFM: free fat mass
BMI: Body mass index

## DECLARATIONS

### Ethics approval and consent to participate

This study was conducted in accordance with the principles of the Declaration of Helsinki. The study protocol was approved by the Makerere University School of Biomedical Sciences Research Ethics Committee, the Uganda National Council on Science and Technology, and the institutional review board at University Hospitals Cleveland Medical Center. Written informed consent was obtained from the parent or guardian of each HHC.

### Consent for publication

Not applicable

### Availability of data and materials

The datasets used and/or analysed during the current study are available from the corresponding author on reasonable request.

### Author Contributions

JG, CMS, and CL contributed to the design and implementation of the research, to the analysis of the results and to the writing of the manuscript, and critically reviewed and revised the manuscript.

JM, MA, SPGM, SA, SB, FA, SK, LM, and EM contributed to the implementation of the research, collection of the data, and critically reviewed and revised the manuscript.

MM contributed to the writing of the manuscript, and critically reviewed and revised the manuscript.

All authors approved the final manuscript as submitted and agree to be accountable for all aspects of the work.

## Acknowledgements

This study would not be possible without the generous participation of the Ugandan children and families. We want to acknowledge the contributions made by home health visitors, clinic providers, data managers, and laboratory personnel: Mary Nsereko, Keith Chervenak, Michael Odie, Hussein Kisingo, Fred Mugaya, Erias Ssaku, and Billy Twaha Mutebi.

## Author’s information

Not applicable

## Footnotes

Not applicable

