## Supplemental materials for "Immune-sensitization to *Mycobacterium tuberculosis* Among Young Children With and Without Tuberculosis"

**SUPPLEMENTAL DATA**

**Supplemental Table 1**: Presence of symptoms and symptom duration of the index cases of study participants with and without TB

| **Symptoms/Symptom duration (days)** | **Index of full cohort** | **Index of PedAS** | **Index of PedTB** | **p-value (test)** |
| --- | --- | --- | --- | --- |
| Cough = present | 130 (100) | 75 (100) | 55 (100) | NA |
| Cough duration | 60 [30-150] | 60 [30-180] | 60 [30-120] | 0.98 (MWU) |
| Fever = present | 76 (65) | 43 (63.2) | 33 (67.3) | 0.66 (χ ^2^) |
| Fever duration | 30 [21-60] | 30 [21-60] | 30 [30-150] | 0.12 (MWU) |
| Productive sputum = present | 114 (97.4) | 68 (100) | 46 (93.9) | 0.14 (χ ^2^) |
| Productive sputum duration | 60 [30-112.5] | 60 [30-97.5] | 60 [30-112.5] | 0.84 (MWU) |
| Purulent sputum = present | 91 (77.8) | 56 (82.4) | 35 (71.4) | 0.24 (χ ^2^) |
| Purulent sputum duration | 60 [30-90] | 60 [30-97.5] | 30 [25.5-90] | 0.25 (MWU) |
| Night sweats = present | 76 (65) | 39 (57.4) | 37 (75.5) | 0.07 (χ ^2^) |
| Night sweats duration | 52.5 [21-90] | 60 [30-90] | 45 [21-90] | 0.94 (MWU) |
| Hemoptysis = present | 14 (12) | 7 (10.3) | 7 (14.3) | 0.71 (χ ^2^) |
| Hemoptysis duration | 6 [3-21] | 21 [4.5-21] | 3 [3-6] | 0.44 (MWU) |
| Weight loss = present | 99 (84.6) | 57 (83.8) | 42 (85.7) | 0.98 (χ ^2^) |
| Weight loss duration | 60 [30-105] | 60 [30-120] | 45 [30-90] | 0.57 (MWU) |

Counts (percentages) or median [quartiles]

PedAS: Not diagnosed with TB

PedTB: Diagnosed with TB

χ ^2^: Chi-squared test

MWU: Mann–Whitney *U* test

**Supplemental Table 2**: Final diagnostic criteria for tuberculosis disease (TB)

**Unconfirmed TB**

Child has a known TB household exposure, plus:

At least one of the following:

- Symptoms/signs suggestive of tuberculosis: cough ≥ 14 days, fever ≥ 7 days, poor appetite, known weight loss or failure to thrive, fatigue and/or reduced playfulness
- Chest x-ray findings consistent with pulmonary TB
- Documented positive response to TB treatment

AND

- Negative microbiological testing

**Confirmed TB**

Positive microbiological testing with or without positive signs and symptoms and chest x-ray consistent with pulmonary TB

**Supplemental Table 3**: Nutritional status among children with and without TB at study entry

| **Z Scores** | **PedAS** | **PedTB** | **p-value (test)** |
| --- | --- | --- | --- |
| n | 78 | 54 |  |
| Weight | -0.72 [+/- 1.18] | -0.41 [+/- 1.18] | 0.14 (t-test) |
| Length/Height | -0.92 [+/- 1.56] | -0.72 [+/- 1.24] | 0.41 (t-test) |
| Weight for Length/Height | -0.29 [+/- 1.28] | -0.03 [+/- 1.10] | 0.21 (t-test) |
| BMI | -0.21 [+/- 1.31] | 0.03 [+/- 1.14] | 0.26 (t-test) |
| Upper arm Circumference | -0.54 [+/- 1.03] | -0.36 [+/- 0.98] | 0.32 (t-test) |

Means [+/- standard deviation]

PedAS: Not diagnosed with TB

PedTB: Diagnosed with TB

BMI: Body mass index

**Supplemental Table 4**: Cross-tabulation of QFT-Plus and TST results (10 mm cutoff) for all participants

|  |  | **TST Results (10 mm cutoff)** | | | |
| --- | --- | --- | --- | --- | --- |
| **Full Cohort** | **QFT-Plus Results** | **N=125*** | **Positive** | **Negative** |  |
|  |  | **Positive** | 37 (29.6%) | 4 (3.2 %) | McNemar χ ^2^ = 0.12 |
|  |  | **Negative** | 11 (8.8%) | 73 (58.4%) | Cohen’s Kappa^a^ = 0.74 |
| **Children <2 years of age** | **QFT-Plus Results** | **N=53** | **Positive** | **Negative** |  |
|  |  | **Positive** | 11 (20.7%) | 1 (1.9 %) | McNemar χ ^2^ = 0.04 |
|  |  | **Negative** | 8 (15.1%) | 33 (62.3%) | Cohen’s Kappa^a^ = 0.60 |
| **Children 2-5 years of age** | **QFT-Plus Results** | **N=72** | **Positive** | **Negative** |  |
|  |  | **Positive** | 26 (36.0%) | 3 (4.2 %) | McNemar χ ^2^ = 1.0 |
|  |  | **Negative** | 3 (4.2%) | 40 (55.6%) | Cohen’s Kappa^a^ = 0.83 |

*Does not include 5 participants with indeterminate QFT results

^a^Cohen’s Kappa interpretation: 0.01–0.20 as none to slight, 0.21–0.40 as fair, 0.41– 0.60 as moderate, 0.61–0.80 as substantial, and 0.81–1.00 as almost perfect agreement.

**Supplemental Table 5**: Cross-tabulation of QFT-Plus and TST results (10 mm cutoff) for PedAS (Not diagnosed with TB) participants

|  |  | **TST Results (10 mm cutoff)** | | | |
| --- | --- | --- | --- | --- | --- |
| **PedAS** | **QFT-Plus Results** | **N=73*** | **Positive** | **Negative** |  |
|  |  | **Positive** | 19 (26%) | 2 (2.7%) | McNemar χ ^2^ = 0.68 |
|  |  | **Negative** | 4 (5.5%) | 48 (65.8%) | Cohen’s Kappa^a^ = 0.80 |
| **Children <2 years of age** | **QFT-Plus Results** | **N=33** | **Positive** | **Negative** |  |
|  |  | **Positive** | 8 (24.3%) | 0 (0%) | McNemar χ ^2^ = 0.13 |
|  |  | **Negative** | 4 (12.1%) | 21 (63.6%) | Cohen’s Kappa^a^ = 0.72 |
| **Children 2-5 years of age** | **QFT-Plus Results** | **N=40** | **Positive** | **Negative** |  |
|  |  | **Positive** | 11 (27.5%) | 2 (5%) | McNemar χ ^2^ = 0.48 |
|  |  | **Negative** | 0 (0%) | 27 (67.5%) | Cohen’s Kappa^a^ = 0.88 |

*Does not include 2 participants with indeterminate QFT results

^a^Cohen’s Kappa interpretation: 0.01–0.20 as none to slight, 0.21–0.40 as fair, 0.41– 0.60 as moderate, 0.61–0.80 as substantial, and 0.81–1.00 as almost perfect agreement.

**Supplemental Table 6**: Cross-tabulation of QFT-Plus and TST results (10 mm cutoff) for PedTB (Diagnosed with TB) participants

|  |  | **TST Results (10 mm cutoff)** | | | |
| --- | --- | --- | --- | --- | --- |
| **PedTB** | **QFT-Plus Results** | **N=52*** | **Positive** | **Negative** |  |
|  |  | **Positive** | 18 (34.6%) | 2 (3.8%) | McNemar χ ^2^ = 0.18 |
|  |  | **Negative** | 7 (13.5%) | 25 (48.1%) | Cohen’s Kappa^a^ = 0.65 |
| **Children <2 years of age** | **QFT-Plus Results** | **N=20** | **Positive** | **Negative** |  |
|  |  | **Positive** | 3 (15%) | 1 (5%) | McNemar χ ^2^ = 0.37 |
|  |  | **Negative** | 4 (20%) | 12 (60%) | Cohen’s Kappa^a^ = 0.39 |
| **Children 2-5 years of age** | **QFT-Plus Results** | **N=32** | **Positive** | **Negative** |  |
|  |  | **Positive** | 15 (46.9%) | 1 (3.1 %) | McNemar χ ^2^ = 0.62 |
|  |  | **Negative** | 3 (9.4%) | 13 (40.6%) | Cohen’s Kappa^a^ = 0.75 |

*Does not include 3 participants with indeterminate QFT results

^a^Cohen’s Kappa interpretation: 0.01–0.20 as none to slight, 0.21–0.40 as fair, 0.41– 0.60 as moderate, 0.61–0.80 as substantial, and 0.81–1.00 as almost perfect agreement.

**Supplemental Table 7**: Cross-tabulation of QFT and TST results (5 mm cutoff) for all participants.

|  |  | **TST Results (5 mm cutoff)** | | | |
| --- | --- | --- | --- | --- | --- |
| **Full Cohort** | **QFT-Plus Results** | **N=125*** | **Positive** | **Negative** |  |
|  |  | **Positive** | 38 (30.4%) | 3 (2.4%) | McNemar χ ^2^ = 0.0003 |
|  |  | **Negative** | 22 (17.6%) | 62 (49.6%) | Cohen’s Kappa^a^ = 0.59 |
| **Children <2 years of age** | **QFT-Plus Results** | **N=53** | **Positive** | **Negative** |  |
|  |  | **Positive** | 11 (20.8%) | 1 (1.9%) | McNemar χ ^2^ = 0.002 |
|  |  | **Negative** | 14 (26.4%) | 27 (50.9%) | Cohen’s Kappa^a^ = 0.42 |
| **Children 2-5 years of age** | **QFT-Plus Results** | **N=72** | **Positive** | **Negative** |  |
|  |  | **Positive** | 27 (37.5%) | 2 (2.8%) | McNemar χ ^2^ = 0.11 |
|  |  | **Negative** | 8 (11.1%) | 35 (48.6%) | Cohen’s Kappa^a^ = 0.72 |

*Does not include 5 participants with indeterminate QFT-Plus results

^a^Cohen’s Kappa interpretation: 0.01–0.20 as none to slight, 0.21–0.40 as fair, 0.41– 0.60 as moderate, 0.61–0.80 as substantial, and 0.81–1.00 as almost perfect agreement.

**Supplemental Table 8**: Cross-tabulation of QFT and TST results (5 mm cutoff) for PedAS (Not diagnosed with TB) participants.

|  |  | **TST Results (5 mm cutoff)** | | | |
| --- | --- | --- | --- | --- | --- |
| **PedAS** | **QFT-Plus Results** | **N=73*** | **Positive** | **Negative** |  |
|  |  | **Positive** | 20 (27.4%) | 1 (1.4%) | McNemar χ ^2^ = 0.03 |
|  |  | **Negative** | 9 (12.3%) | 43 (58.9%) | Cohen’s Kappa^a^ = 0.70 |
| **Children <2 years of age** | **QFT-Plus Results** | **N=33** | **Positive** | **Negative** |  |
|  |  | **Positive** | 8 (24.2%) | 0 (0%) | McNemar χ ^2^ = 0.02 |
|  |  | **Negative** | 7 (21.2%) | 18 (54.6%) | Cohen’s Kappa^a^ = 0.55 |
| **Children 2-5 years of age** | **QFT-Plus Results** | **N=40** | **Positive** | **Negative** |  |
|  |  | **Positive** | 12 (30%) | 1 (2.5%) | McNemar χ ^2^ = 1 |
|  |  | **Negative** | 2 (5%) | 25 (62.5%) | Cohen’s Kappa^a^ = 0.83 |

*Does not include 2 participants with indeterminate QFT results

^a^Cohen’s Kappa interpretation: 0.01–0.20 as none to slight, 0.21–0.40 as fair, 0.41– 0.60 as moderate, 0.61–0.80 as substantial, and 0.81–1.00 as almost perfect agreement.

**Supplemental Table 9**: Cross-tabulation of QFT and TST results (5 mm cutoff) for PedTB (Diagnosed with TB) participants.

|  |  | **TST Results (5 mm cutoff)** | | | |
| --- | --- | --- | --- | --- | --- |
| **PedTB** | **QFT-Plus Results** | **N=52*** | **Positive** | **Negative** |  |
|  |  | **Positive** | 18 (34.6%) | 2 (3.9%) | McNemar χ ^2^ = 0.01 |
|  |  | **Negative** | 13 (25%) | 19 (36.5%) | Cohen’s Kappa^a^ = 0.45 |
| **Children <2 years of age** | **QFT-Plus Results** | **N=20** | **Positive** | **Negative** |  |
|  |  | **Positive** | 3 (15%) | 1 (5%) | McNemar χ ^2^ = 0.08 |
|  |  | **Negative** | 7 (35%) | 9 (45%) | Cohen’s Kappa^a^ = 0.20 |
| **Children 2-5 years of age** | **QFT-Plus Results** | **N=32** | **Positive** | **Negative** |  |
|  |  | **Positive** | 15 (46.9%) | 1 (3%) | McNemar χ ^2^ = 0.13 |
|  |  | **Negative** | 6 (18.8%) | 10 (31.3%) | Cohen’s Kappa^a^ = 0.56 |

*Does not include 3 participants with indeterminate QFT results

^a^Cohen’s Kappa interpretation: 0.01–0.20 as none to slight, 0.21–0.40 as fair, 0.41– 0.60 as moderate, 0.61–0.80 as substantial, and 0.81–1.00 as almost perfect agreement.

**Supplemental Table 10:** Logistic regression model of predictors of PedTB (Symptomatic for TB) adjusted for age (months), sex, HIV status, and BCG status.

| **COVARIATES** | **ADJUSTED OR** | **95% CI** | ***p*-VALUE** |
| --- | --- | --- | --- |
| Quantitative TST result (in mm) | 1.04 | 1.001 – 1.09 | 0.051* |
| Sex = male | 0.90 | 0.42 – 1.91 | 0.78 |
| Age (months) | 1.01 | 0.99 – 1.04 | 0.21 |
| HIV = positive | 0.91 | 0.04 – 10.42 | 0.94 |
| BCG scar = present | 1.25 | 0.44 – 3.77 | 0.68 |

*** Statistically significant at *p*<0.05**

CI: Confidence interval

**Supplemental Table 11**: Quantitative IGRA results summary (n=125*) by Tube

|  | **Minimum** | **First quartile** | **Median** | **Third quartile** | **Maximum** | **Mean** | **Standard deviation** |
| --- | --- | --- | --- | --- | --- | --- | --- |
| **Tube TB.1** | 0 | 0 | 0.01 | 1.85 | 10 | 1.94 | 3.39 |
| **Tube TB.2** | 0 | 0 | 0.02 | 2.78 | 10 | 1.99 | 3.38 |

*Does not include five participants with indeterminate results

**Supplemental Table 12**: Quantitative IGRA results among 3 participants with discordant QFT-Plus Tube TB.1 minus nil and Tube TB.2 minus mil results

| **Age group** | **Tube TB.1** | **Tube TB.2** |
| --- | --- | --- |
| Older than 2 years | 0.02 IU | 3.01 IU |
| Older than 2 years | 0.34 IU | 0.55 IU |
| Older than 2 years | 0.97 IU | 0.00 IU |

**Supplemental Figure 1**: Form used for the evaluation of chest x-rays obtained during the study

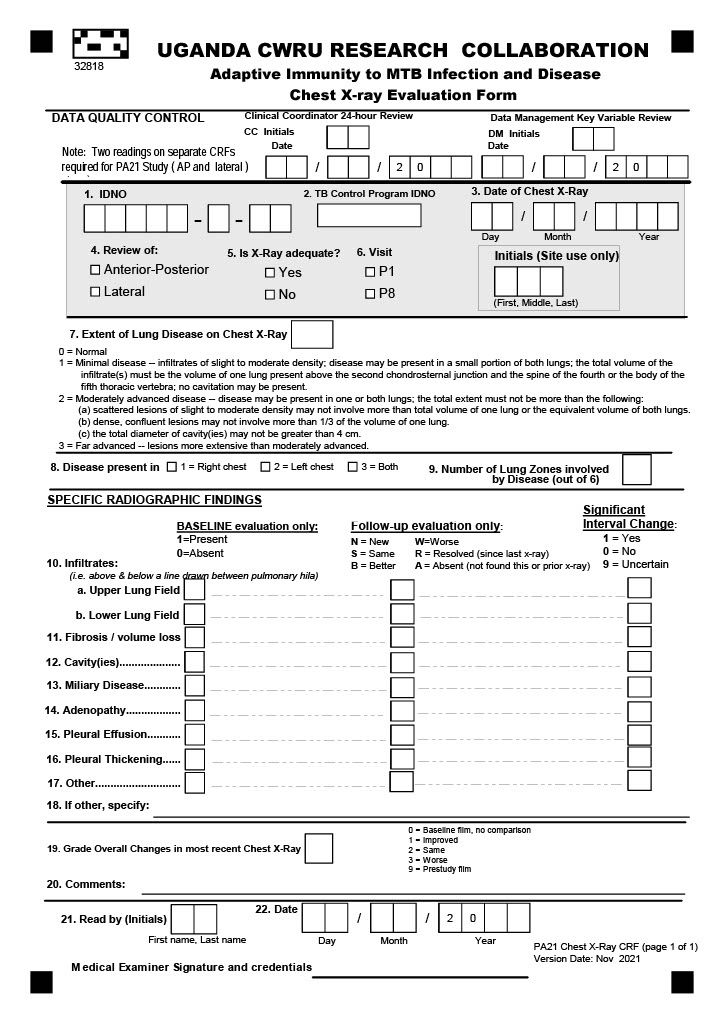
